# Feasibility of an Evolutionary Tumor Board for Generating Novel Personalized Therapeutic Strategies

**DOI:** 10.1101/2023.01.18.23284628

**Authors:** Mark Robertson-Tessi, Joel S. Brown, Maria I. Poole, Matthew Johnson, Andriy Marusyk, Jill A. Gallaher, Kimberly A. Luddy, Christopher J. Whelan, Jeffrey West, Maximillian Strobl, Virginia Turati, Heiko Enderling, Michael J. Schell, AikChoon Tan, Terry Boyle, Rikesh Makanji, Joaquim Farinhas, Hatem Soliman, Dawn Lemanne, Robert A. Gatenby, Damon R. Reed, Alexander R. A. Anderson, Christine H. Chung

**Affiliations:** Department of Integrated Mathematical Oncology, H. Lee Moffitt Cancer Center and Research Institute, Tampa, Florida; Department of Head and Neck-Endocrine Oncology, H. Lee Moffitt Cancer Center and Research Institute, Tampa, Florida; Department of Cancer Physiology, H. Lee Moffitt Cancer Center and Research Institute, Tampa, Florida; Department of Biostatistics and Bioinformatics, H. Lee Moffitt Cancer Center and Research Institute, Tampa, Florida; Department of Pathology, H. Lee Moffitt Cancer Center and Research Institute, Tampa, Florida; Department of Radiology, H. Lee Moffitt Cancer Center and Research Institute, Tampa, Florida; Department of Breast Oncology, H. Lee Moffitt Cancer Center and Research Institute, Tampa, Florida; Department of Individualized Cancer Management; H. Lee Moffitt Cancer Center and Research Institute, Tampa, Florida; Department of Oncological Sciences, Huntsman Cancer Institute, University of Utah, Salt Lake City, UT; Oregon Integrative Oncology, Ashland, Oregon; Department of Medicine, University of Arizona, Tucson, Arizona

**Keywords:** evolution, multi-disciplinary cancer care, adaptive therapy, mathematical models, evolutionary therapy

## Abstract

The current paradigm of clinical trials treating patients until disease progression using maximum tolerated dose does not account for the dynamic tumor-host-drug interactions that result in acquired resistance. Here, we present the concept of an Evolutionary Tumor Board (ETB) and report interim results from a prospective, non-interventional pilot study in which novel therapeutic strategies based on evolutionary principles were developed under the ETB framework. The ETB approach relies on an interdisciplinary team that integrates clinical, preclinical, and theoretical knowledge and the application of mathematical modeling to predict patient responses to different therapies, including novel approaches derived from eco-evolutionary first principles. We have previously proposed several evolutionary therapies that aim to enhance the efficacy of an overall treatment regimen, using existing agents for a given disease. Key among these evolutionary therapies is the idea of “first-strike second-strike”, where different agents are administered in sequence, and new strikes are applied as soon as the efficacy of the previous strike is nearing a minimum, as opposed to waiting until progression is identified on periodic imaging. This approach requires careful analysis of longitudinal patient data coupled with predictive dynamics generated by mathematical models. Here we describe the ETB process and the interim results from 15 patients enrolled in the feasibility trial. In addition, we describe the challenges faced as well as the solutions that can be implemented via improved modeling approaches, better patient data collection, and a reassessment of how we understand tumor dynamics in the light of evolutionary principles.

## INTRODUCTION

Cancer is one of the major public health concerns globally, and the American Cancer Society estimates approximately 1,918,030 new cases will be diagnosed in the United States alone in 2022^1^. With breakthrough advances in cancer screening, targeted therapies, and immunotherapies, overall cancer mortalities have been decreasing. However, cancer is still the second leading cause of death in the U.S., with an estimated 609,360 people expected to die from cancer in 2022. Therefore, the development of novel strategies to improve outcomes remains urgent.

The general paradigm of novel therapy development in currently “incurable” cancers (by current standard of care) that are recurrent and/or metastatic with resistance to existing treatments has been focused on molecular characterization of a tumor for targeted therapies, leveraging immune checkpoint inhibition, and identifying predictive biomarkers to select the most efficacious regimen. However, this approach benefits only a limited number of patients with targetable genomic aberrations and tumors with immune responsive phenotypes; furthermore, most of these patients exhibit transient responses due to the evolution of acquired resistance over time. This limitation is, at least in part, due to the lack of a comprehensive evaluation of the dynamic tumor-host-treatment interactions over time. Consideration of evolution under treatment selection and subsequent changes in tumor phenotype is not a novel concept; nevertheless, strategies for using this perspective to guide systemic therapies are lacking. Too often oncology strategies are static. The prevailing clinical intervention is reactionary after resistance develops, and not proactive to curtail evolutionary dynamics^2^. From the development of the first cancer cell, natural selection favors phenotypes that increase fitness and eliminates those that do not, and this process continues through therapy^3,4^. We continue to select systemic therapies empirically based on the observable response in the dominant population because we are often unable to account for resistant minor populations during the treatments that will eventually lead to recurrence. For patients with incurable cancers, we must better understand the evolutionary dynamics that lead to the emergence and predominance of therapy-resistant phenotypes, eventually rendering even initially useful therapies to be ultimately ineffective. This highlights our currently inability to incorporate evolutionary dynamics into routine clinical treatment to delay or avoid the emergence of treatment resistance. But what is a better way?

There are several concepts from evolution and ecology that can potentially address these limitations. For example, Anthropocene species extinctions have commonly occurred when a cataclysmic perturbation (first strike) spatially fragments a large population into small remnant populations with less genetic diversity and increased sensitivity to stochasticity that then become susceptible to what otherwise might be survivable minor perturbations (second strikes, third strikes, etc.). This concept could be leveraged to develop an “extinction treatment” regimen that aims to improve therapy by using a “first-strike” agent to generate a “cataclysmic” response, followed by a sequence of second-strike therapy agents that might offer patients the chance of cure or complete response^5^. Furthermore, even if cure is not possible with such a sequence of strikes, moving agents forward to apply them near the nadir of the previous strike has the potential to foment greater overall tumor decline, and the dynamics of these diminished tumor burdens may prolong survival.

The vast majority of currently employed strategies against cancer can be considered as first strikes. Practically speaking, first strikes are any intervention such as surgery, radiation, or systemic therapy agent(s) that induce an observable response. Current first strikes are evaluated by how large a response occurs (with partial or complete responses seen as desirable) and how durable the response remains before progression. In the current fixed maximum tolerated dose paradigm, the subsequent therapy is not initiated until the observation of clear disease progression after the first strike. However, changing therapies upon disease progression is far too late in eco-evolutionary terms. Clinical progression simply provides delayed proof of the evolutionary and ecological recovery of cancer cell populations, often weeks or even months (or years or more) after the fact. Instead, the second strike should occur when, or even before, the cancer cell population is the smallest after the first strike. Second-strike therapies do not need to have a proven track record for producing a large tumor response like first-strike agents. Importantly, any remnant cancer populations survived by the first strike may have vulnerabilities (i.e., collateral sensitivity) to second-strike agents^5,6^.

Application of extinction-based therapies poses two questions to practicing clinicians: 1) what is the appropriate timing of the second strike? and 2) what therapeutic agent(s) should be selected as the second strike? To address these fundamental questions, we adapted the existing framework of the clinical tumor board that determines the best treatment options using multidisciplinary approaches and formed the Evolutionary Tumor Board (ETB), broadening the traditional tumor board to include the non-clinical disciplines such as Cancer Biology, Evolutionary Biology, Mathematical Oncology, and Bioinformatics. The ETB explicitly considers the ecological (changes in tumor burden and distribution of tumors) and evolutionary (changes in the heritable characteristics of the cancer cell populations) dynamics of cancer. We hypothesize that this ETB approach can develop novel therapeutic strategies based on eco-evolutionary principles that may provide longer lasting responses by using currently available drugs in new ways.

At the heart of evolutionarily inspired cancer therapies, cancer is seen as a complex, adaptive dynamic system governed by natural selection. Evolutionary therapies model cancer cells within a tumor as an adapting and phenotypically heterogeneous population. Central to capturing cancers’ complex dynamics is patient-specific mathematical modelling (e.g. using differential equations or game theory), with its ability to exploit historic response data (for model calibration) and forecast future therapy combinations and schedules^7^. Integrating clinical experience, completed clinical trial data, retrospective patient data analysis, extensive literature review, and detailed patient specific analysis facilitates a unique dialogue among the many involved disciplines. This results in a patient-specific decision tool, driven by predictive mathematical modeling, for evaluating the possible consequences of different treatment options. Importantly, this approach allows for detailed reassessment of care decisions as follow-up data is received during treatment. Here, we report a description of the ETB workflow and framework, its feasibility, and the interim results of this pilot study.

## METHODS

### Study Design and Patient Selection

The prospective, non-interventional pilot study was conducted at Moffitt Cancer Center under an IRB-approved protocol (MCC 20417, Feasibility of Generating Novel Therapeutic Strategies based on Evolutionary Tumor Board, NCT04343365). Institutional IRB approval was obtained in accordance with the Department of Health and Human Services Federal Policy for the Protection of Human Subjects (US Common Rule) at Moffitt Cancer Center. The study was initiated after the IRB approvals and written consents were obtained. Patients were eligible for enrollment if they were deemed to be incurable given the current standards of care, had a life expectancy over 3 months, an Eastern Cooperative Oncology Group (ECOG) performance status of 0-2, had a primary oncologist willing to consider the therapeutic strategies recommended by the ETB, and were willing to be followed over time and allowing clinical data collection over time. This includes patients in remission but at high risk of recurrence, patients with suboptimal responses to previous therapy, or patients with many potentially beneficial (but not curative) options for care. There were no exclusion criteria.

The primary endpoint of the study is to determine the rate of developing evolutionary-therapy-based treatment strategies for ETB-enrolled patients without curative options. Exploratory aims include other potential results of this process: 1) To determine the rate of implementing evolution-inspired plans by the treating physician through a chart review; 2) To assess whether evolutionary strategies recommended by the ETB improve prognosis compared to *a priori* prognosis for patients who have exhausted curative strategies through a chart review; 3) To assess the feasibility to build and refine mathematical models to explore duration of effect and survival for ETB recommendations; 4) To assess the feasibility to analyze radiologic features of cancer over time to predict duration of effect and survival for ETB recommendations; 5) To assess whether changes in the circulating cell-free DNA (cfDNA) over time as a variable is or is not informative in building mathematical models; 6) To assess whether presence of certain tumor genomic abnormalities by whole exome sequencing as a variable is or is not informative in building mathematical models; and 7) To assess the feasibility of generating impact hypotheses based on evolutionary and/or ecological principles to improved cancer therapy.

### The ETB Process

Once enrolled, a patient’s disease history and treatment summary were compiled by the oncologist and the ETB coordinator. The remaining options for care and anticipated response rates along with prognosis in terms of progression-free survival (PFS) and overall survival (OS) were annotated from the literature. Collected data points include 1) demographics, 2) social history including alcohol and tobacco use history, 3) family history, 4) Eastern Cooperative Oncology Group performance status, 5) pathology, 6) any clinically relevant laboratory test and biomarker results, 7) prior therapies and associated tumor measurements during the therapies, and 8) potentially available subsequent therapies for consideration by the ETB. The tumor measurements were obtained as 1-dimensional measurements using the RECIST v1.1 criteria as a guideline and 3-dimensional measurements for volumetric quantification of all measurable lesions. In addition, for the purpose of calibrating the model, we typically utilize retrospective clinical data for the drugs under consideration for a given patient. For the specific exemplar patient below, we used data from a phase II study of cetuximab and nivolumab in recurrent and/or metastatic (R/M) head and neck squamous cell carcinoma (HNSCC; NCT03370276). The detailed study population and results were previously published^8,9^.

For each patient, at least 2 preparatory discussion meetings occurred among a subset of the key ETB members in advance to the ETB meeting, consisting at least of primary oncologists that oversee the treatment implementation of the patient being discussed, mathematical modelers, preclinical experimentalists, ecologists, radiologists, and often other oncology disciplines and additional *ad hoc* members. During these discussions, the current clinical standard was discussed along with alternatives. Evolutionary strategies were proposed to establish what interventions were possible and what measurements of responses were feasible. Predictions from a patient-calibrated mathematical model and several iterations of the clinical questions in response to them often occurred during the generation of the ETB recommended treatment plan.

The ETB itself was convened monthly and consisted of clinical oncologists (surgical, radiation, medical, and pediatric), radiologists, pathologists, evolutionary biologists, mathematicians, research scientists, statisticians, data scientists, and clinical trial-related personnel at Moffitt Cancer Center. The primary oncologist typically started by presenting their case to the multidisciplinary ETB, followed by the mathematical modelling team presenting the process and initial thoughts and plans for the patient. The interdisciplinary discussion that followed often refined and sometimes changed a therapeutic strategy, and also generated hypotheses that influenced directions for follow up preclinical work, future clinical trials, or other areas of investigation.

## RESULTS

### Patient Characteristics

A total of 15 patients were enrolled to the ETB protocol between 5/5/2020 and 4/7/2022. As described below, we also used a retrospective cohort to calibrate the model. This cohort consisted of a total of 26 patients with HNSCC and were obtained from the phase II clinical trial of cetuximab and nivolumab^8,9^. The patient characteristics for the ETB and retrospective patients are summarized in **Table 1**. The prior line of therapy is defined as the number of treatment regimens that the patient received from the time of recurrent and/or metastatic disease diagnosis. Most patients were heavily treated before enrolling to the trial, but 14 of 15 (93%) patients had good performance status with ECOG PS 0-1.

**Table 1.** Patient characteristics.

| **Variables** |  | **ETB**<br>**N=15** | **HNSCC**<br>**Retrospective**<br>**Cohort**<br>**N=26** |
| --- | --- | --- | --- |
| **Age** | Median | 59 | 64 |
| **Gender** | Male | 12 (80%) | 23 (88%) |
|  | Female | 3 (20%) | 3 (12%) |
| **Race** | White | 13 (87%) | 25 (96%) |
|  | Black | 1 (7%) | 0 |
|  | Other | 1 (7%) | 1 (4%) |
| **ECOG PS** | 0 | 8 (53%) | 7 (27%) |
|  | 1 | 6 (40%) | 19 (73%) |
|  | 2 | 1 (7%) | 0 |
| **Smoking**<br>**History** | Yes | 6 (40%) | 17 (65%) |
|  | No | 9 | 9 |
| **Disease Site** | HNSCC | 6 (40%) | 26 (100%) |
|  | Sarcoma | 7 (47%) | 0 |
|  | Lung | 1 (7%) | 0 |
|  | Prostate | 1 (7%) | 0 |
| **Prior line of**<br>**therapy for R/M**<br>**cancers** | 0 | 0 (0%) | 6 (23%) |
|  | 1 | 6 (40%) | 13 (50%) |
|  | 2 | 6 (40%) | 5 (19%) |
|  | 3 | 0 (0%) | 2 (8%) |
|  | 4 | 3 (20%) | 0 (0%) |

ETB: evolutionary tumor board
HNSCC: head and neck squamous cell carcinoma
ECOG PS: Eastern Cooperative Oncology Group Performance Status
R/M: recurrent and/or metastatic

### Development of Therapeutic Strategies based on Mathematical Modeling

The overall ETB workflow is summarized in **Figure 1A**. Each individual patient’s history was collected based on the protocol requirement and summarized. Examples of the prior treatment history and data collection along with available treatment options and their estimated outcome are shown in **Supplemental Table 1** and **Supplemental Table 2**. The collected data were graphed to visualize an accurate timeline of disease burden based on the radiographical imaging studies, drug dosing, and all sequences of previous treatments (**Figure 1B**). If imaging data informed tumor burden, then the volume dynamics, appearance, and disappearance of each lesion were also included. Aggregating this information into a standardized visual treatment and response chart has proved to be an invaluable tool for ETB discussion.

**Figure 1.**
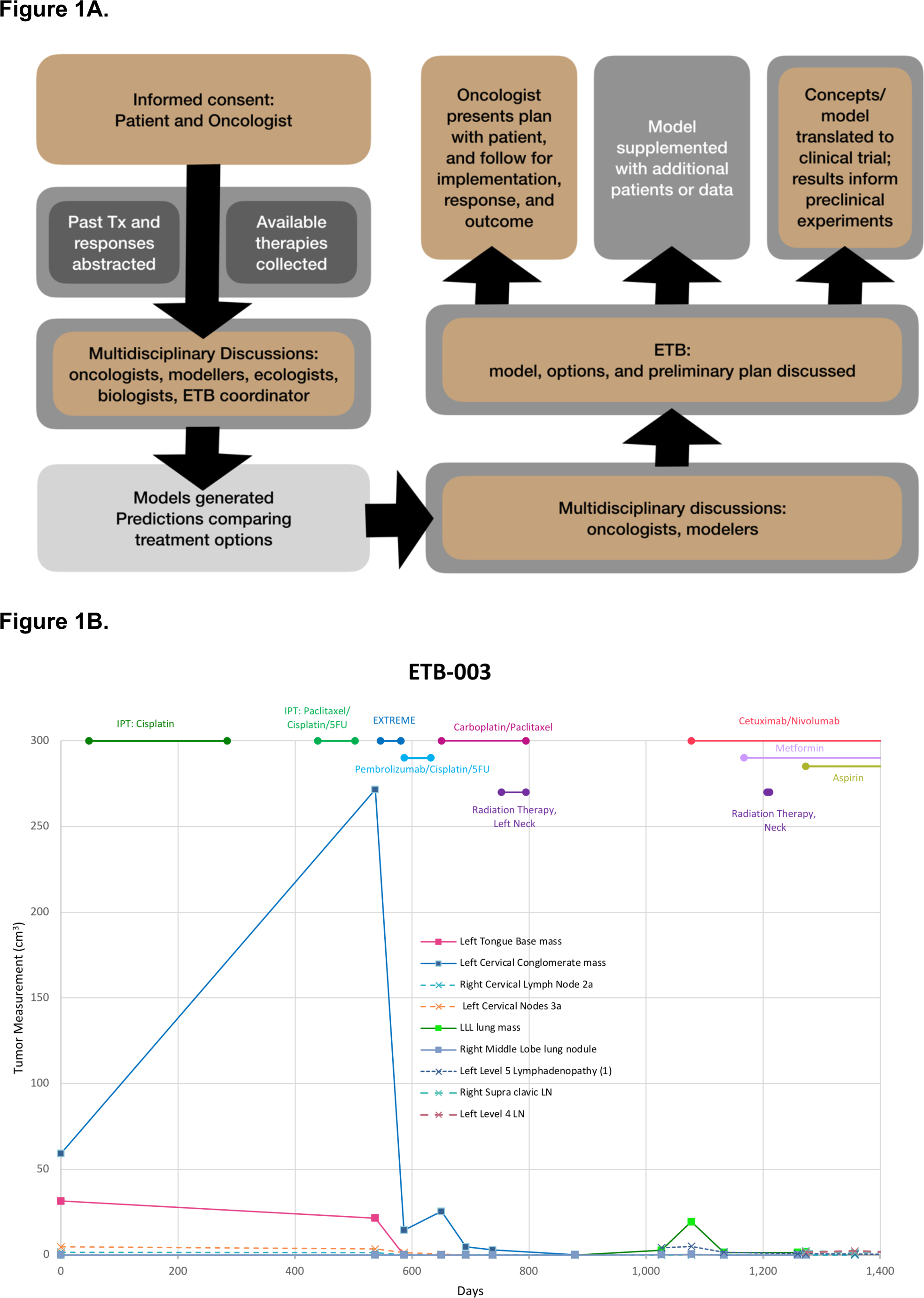
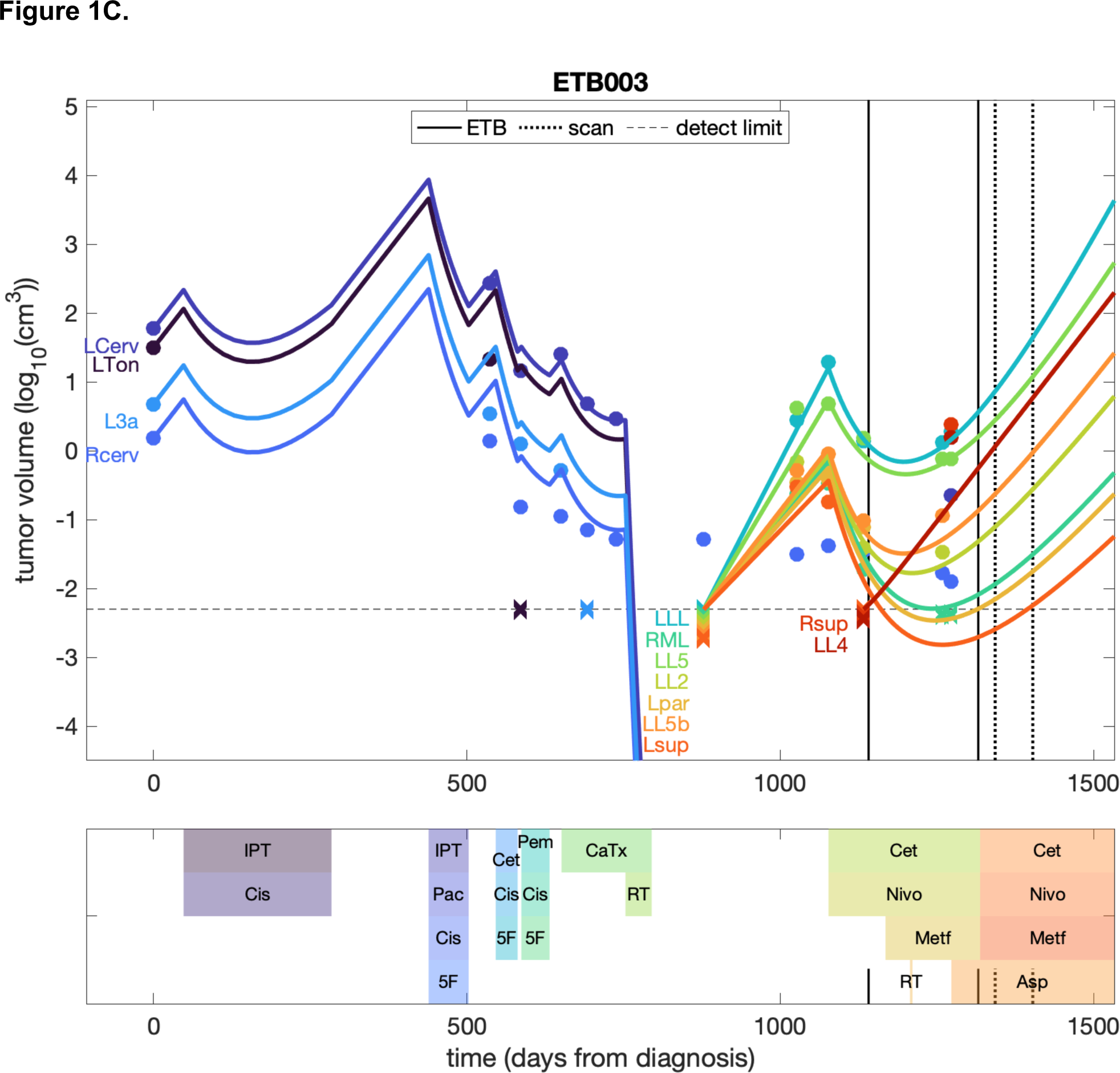
**A:** Schematic diagram of the Evolutionary Tumor Board clinical workflow. **B:** An example of tumor volumetric data visualization in context of the clinical data. **C:** An example of tumor volumetric data visualization based on a mathematical model using a system of ordinary differential equations in context of the treatment data.

The available clinical data points were integrated to calibrate mathematical models that explore treatment options using a system of ordinary differential equations (ODEs). The equations have sufficient complexity to capture the key disease dynamics observed in patients and remain simple enough to avoid overfitting (**Figure 1C**). Data available for each individual patient tend to be sparse in terms of the number of time points, and thus unsuitable for fitting models with numerous parameters^10^. In our primary model, termed the GDRS model, we focus on four aspects of tumor dynamics: tumor <u>G</u>rowth, tumor <u>D</u>eath, evolution of drug <u>R</u>esistance, and drug re-<u>S</u>ensitization. The model is an extension of a tumor-growth inhibition model^11-14^ and consists of *n+m* differential equations, where *n* is the number of distinct lesions, and *m* the number of drugs that are administered. We let *T*_*i*_ be the volume of lesion *i*=1,…*n*; and *D*_*j*_ be the dose (as a function of time) of drug *j=*1,*…,m*. The efficacy of each drug over time (*E*_*j*_) is distinct for each drug, and changes to reflect the evolution of resistance or subsequent resensitization. In the following equations, *T*_*i*_, *D*_*j*_ and *E*_*j*_ are time-dependent:

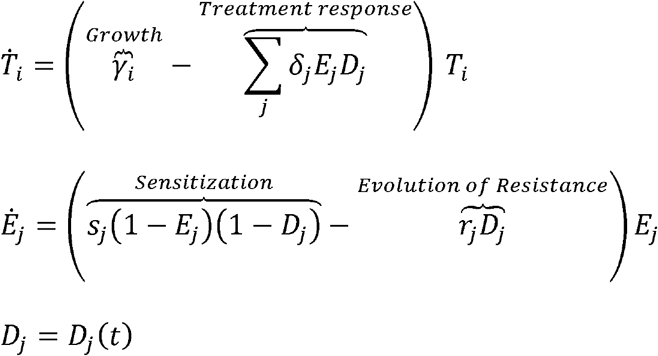

In this eco-evolutionary model, *T*_i_ models the ecological dynamics, *E*_j_ the evolutionary dynamics, and *D*_j_ the choices of the physician and patient. The model is further described in **Supplemental File 1**.

### Model Calibration using ETB Patient Specific Data

To apply the model to an ETB patient’s data, we made the following assumptions. First, we assumed that all the lesions within a patient shared the same growth parameter (γ_*i*_) unless there were significant indications from the data suggesting that a particular lesion should have its own individualized fit. Thus, tumors within a patient only differed in terms of their date of first appearance and initial size. When possible, growth rates were calculated from a pair of consecutive increasing volumetric measurements for a lesion. These pairs of points were picked with the following prioritization scheme: 1) two measurements for the primary lesion prior to initial therapy; 2) two measurements for any metastatic lesion while off therapy; 3) maximal growth rate found within all increasing sequential measured volumetric pairs regardless of therapy status. In cases where multiple pairs of points satisfied the same level from the prioritization scheme, the maximal growth rate was used as the baseline. The growth rate, γ, thus derived is set to be the exponential growth parameter for the patient as a whole, for all lesions. An exception arises if one or more lesions are clearly significantly different in its growth rates, in which case the outlier lesion is given its own growth rate γ_*i*_.

An additional factor can affect the growth rates, namely the appearance of new lesions relative to the dates of patient scans. When a new lesion is detected, the volume is noted. Following that, the previous scan of that same area is rechecked for any prior evidence of that lesion. This is a relevant check as the threshold for identifying a lesion *de novo* is larger than identifying it *post hoc*, when the location is now known from later scans. In either case, imaging has a lower limit of measurable size determined by contrast, voxel size, etc. When tracing a new lesion back through previous scans, eventually a scan is reached where there is no evidence of that lesion. For the model, we set its initial size in accordance with the minimal detection size of the instrument. Thus, this may be an over-estimate and consequently may underestimate the tumor’s growth rate when calculated from the next scan, where the lesion was identified and measured. In some cases, this underestimate may still be higher than the growth rates measured from other lesions, in which case the new lesion may receive its own higher growth rate for fitting purposes. This approach of tracking individual lesions backwards in time is a novel aspect of the ETB, allowing for the extraction of additional data regarding the lesion dynamics that would otherwise not be available.

To fully fit the drug-induced death and resistance parameters, at least two volumetric measurements while on the same therapy are needed. In this case, the starting tumor size (calculated from the pre-treatment scan), the growth rate, and the two on-treatment time points are sufficient to fit the ‘U-shaped’ ecological dynamics of the tumors. The model then reflects initial drug efficacy (drug-induced death rate, δ) followed by increasing drug resistance. In cases where there is only one on-treatment measurement, drug-induced death rate and rate of increasing drug resistance, *r*, cannot both be estimated. In such cases, we specify a functional form that generates pairs of parameters that fit the single on-treatment data point (**Supplemental File 1**). This function defines a set of parameter pairs all of which fit the patient data points. In principle, this set is wide-ranging, although biologically realistic bounds can be placed on δ and *r*, particularly using historical data from independent cohorts. The predictions arising from the possible pairs of δ and *r* can vary greatly. Thus, constraining the set of plausible pairs is of high value for predicting the future course of the patient’s disease.

### Model Calibration using Historical Data

A key aspect of the ETB process is the use of retrospective cohort data to constrain the predictions generated for the specific ETB patient. As such, we evaluated a retrospective cohort of patients with recurrent and/or metastatic HNSCC (**Tables 1 and 2**). For these 26 retrospective patients, we applied the ETB analysis approach to their available historical data. Imaging scans were retrieved and remeasured to both generate volumetric measures of each lesion and look for non-target lesions that may not have been considered during typical RECIST follow-up analysis at the time of their care. These data were then modeled using the above procedures to find parameter ranges for each patient that matched their data. Specifically, ranges of growth rates for lesions (γ), their response to any applied therapies (δ), and their rate of becoming resistant to such therapies (*r*) were determined. These ranges give a starting point for predicting the outcomes of the ETB patient, putting expected bounds on their growth rate and response to treatments, wherever the ETB patient’s data itself does not offer a fit. In our exemplar patient that we describe below, we used the retrospective cohort of patients receiving combination cetuximab and nivolumab to predict the widest range of response to these agents expected in the ETB patient. The patient’s own lesion dynamics then further refines the predictions within that retrospective range of possibilities.

**Table 2.**
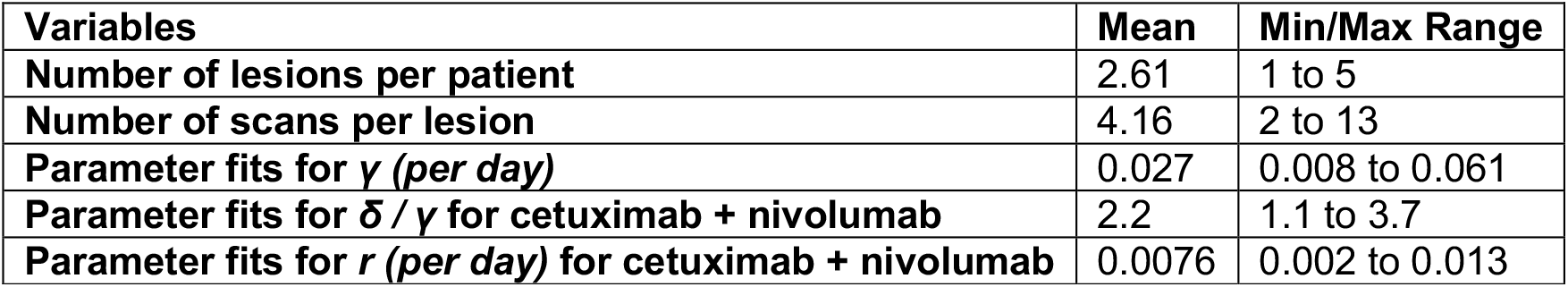
Retrospective head and neck squamous cell carcinoma cohort characteristics (N=26)

| **Variables** | **Mean** | **Min/Max Range** |
| --- | --- | --- |
| **Number of lesions per patient** | 2.61 | 1 to 5 |
| **Number of scans per lesion** | 4.16 | 2 to 13 |
| **Parameter fits for $\gamma$ (per day)** | 0.027 | 0.008 to 0.061 |
| **Parameter fits for $\delta / \gamma$ for cetuximab + nivolumab** | 2.2 | 1.1 to 3.7 |
| **Parameter fits for $r$ (per day) for cetuximab + nivolumab** | 0.0076 | 0.002 to 0.013 |

### Evaluation of Primary Endpoint of the Pilot Study

In 15 patients enrolled to date, 11 patients (73%) met the primary endpoint (**Table 3**). In cases where the end point was not met, the reasons are: 1) the patient was deceased before the date of the first ETB presentation, 2) insufficient historical data/analysis at the time of ETB to predict response to additional therapy options, 3) the patient was taken off the study at physician’s discretion before the ETB presentation, and 4) the patient did not have measurable lesions delaying the presentation at ETB. In the 11 cases with recommendations, subsequent systematic chart reviews were conducted to determine if the treating physician altered the patient’s treatment plan based on the evolutionary therapies recommended by the ETB. The physician and patient followed the recommendation of the model in all 11 cases. All patients were longitudinally followed on the protocol for continued chart review to evaluate, after sufficient follow up, whether the patient had an improved outcome compared to the *a priori* prognosis for similar patients under standard of care.

**Table 3.** Evolutionary Tumor Board (ETB) Recommendations.

| ETB ID | Clinical Presentation at ETB | ETB Recommendation | Implemented by physician |
| --- | --- | --- | --- |
| ETB-001 | Metastatic Ewing sarcoma with prolonged stable disease and unresectable disease burden | Three strike strategy: Short course doxorubicin. Radiation therapy to lung metastasis (whole lung vs stereotactic depending on response) with a systemic therapy backbone (tyrosine kinase inhibitor or vincristine) and then return to vincristine and irinotecan cycled with a tyrosine kinase inhibitor (regorafenib). | doxorubicin + dexrazoxane + cyclophosphamide |
| ETB-002 | Metastatic alveolar rhabdomyosarcoma entering second complete response | Radiation therapy to the forearm lesion, chemotherapy, anti-angiogenesis inhibitor (rotating 90-day cycles) | pazopanib + radiation therapy |
| ETB-003 | Metastatic HPV positive poorly differentiated carcinoma of base of tongue who presented while on nivolumab + cetuximab clinical trial | Maintain combination of nivolumab + cetuximab treatment for at least 90 days from initiation, but not greater than 120 days based on the model. Then switch to a second strike with an available chemotherapy with nivolumab. Nivolumab will be continued as maintenance. Maintain second strike for 90-120 days and then consider switching the chemotherapy again based on the current estimated tumor proliferation rate. | continued nivolumab + cetuximab |
| ETB-004 | Metastatic HPV positive base of tongue squamous cell carcinoma presented with progressive disease on the second line palliative systemic therapy | Discontinue current therapy and initiate concurrent cisplatin or carboplatin + paclitaxel with radiation as a second strike to achieve durable locoregional control. Possibility of radiating the sternal metastasis for palliation will be assessed by the radiation oncology team. As soon as the patient recovers from the radiation induced toxicities, resume an appropriate systemic therapy again to control the lung metastasis. | cisplatin + radiation therapy |
| ETB-005 | Widespread metastatic bone and lung osteosarcoma in first 6 months of MAP | No: Recommendation N/A due to death of the patient after enrolling to the study but before the ETB presentation. | N/A |
| ETB-006 | p16-positive right tonsil squamous cell carcinoma progressed on the first line of palliative systemic therapy | Discontinue the first line carboplatin + paclitaxel. Switch to cisplatin + 5-fluorouracil + pembrolizumab as a second strike. Continue pembrolizumab as maintenance. Introduce second strike of a dose-reduced cisplatin + 5-fluorouracil because the patient had problems with neutropenia given the prior regimen. Alternately, cisplatin + 5-fluorouracil can be given as a full dose by adding a growth factor to prevent prolonged neutropenia. | cisplatin + 5-fluorouracil + pembrolizumab |
| **ETB-007** | Gleason 8 Prostate cancer with widespread bone metastases | Begin an adaptive therapy using enzalutamide + relugolix. Use 50% PSA decline as threshold for stopping therapy and increase of PSA to resume. Follow patient closely to calibrate modeling and potentially alter the recommendation of start-stop algorithm | enzalutamide + relugolix |
| **ETB-008** | Osteosarcoma with metastases in lung with good response | No: Recommendation N/A due to insufficient historical data/analysis at the time of ETB to predict response to additional therapy options. | N/A |
| **ETB-009** | Widely metastatic embryonal rhabdomyosarcoma with LN, bone and lung metastases newly diagnosed | If the first strike results in disease progression or stable disease, then change to doxorubicin-based therapy. If partial response after the first evaluation, continue vincristine + Adriamycin + cyclophosphamide (VAC) and determine intensify or de-intensify the regimen. Second strike: vincristine + irinotecan after complete response between 12-42 weeks of VAC. | VAC → vincristine + irinotecan |
| **ETB010** | Metastatic HPV positive base of tongue squamous cell carcinoma | Await the next follow-up scan to refine the model predictions and decide when to switch to next line of therapy. | pembrolizumab |
| **ETB-011** | Metastatic oropharynx (posterior pharyngeal wall) squamous cell carcinoma | Switch to cetuximab + pembrolizumab instead of administering the final cycle of cisplatin + 5-fluorouracil + pembrolizumab based on evidence of progression. Expect cetuximab to provide 2 to 7 months of efficacy. Nadir of efficacy expected in 1 to 4 months, so close attention to upcoming scans should be given to anticipate the next switch of therapy. | cetuximab + pembrolizumab |
| **ETB-012** | Adenocarcinoma with metastasis to liver | No: Recommendation N/A due to the patient taken off the study at physician's discretion before the ETB presentation. | N/A |
| **ETB-013** | Synchronous floor of mouth (cT1N0M0) and left base of tongue (cT2N0M0) squamous cell carcinoma, both p16 negative. | Next scan will be critical for predicting nadir of response. Scan delayed by insurance, so results pending for now. Consider starting the palliative radiation therapy sooner after getting the next scan. | palliative radiation therapy |
| **ETB-014** | Recurrent esthesioneuroblastoma | No: Recommendation N/A due to the lack of measurable lesions and delay in patient presentation. | N/A |
| **ETB-015** | Bone metastatic Ewing sarcoma with initial response to chemotherapy | Continue vincristine + doxorubicin + cyclophosphamide /ifosfamide + etoposide (VDC/IE). Patient is in the 2nd strike window. | continue VDC/IE |

The ETB has developed a framework to evaluate novel therapeutic strategies for individual patients, including tools for temporal visualization of the treatment and responses throughout the patient’s cancer journey, and application of the GDRS model to volumetric and other biomarker data. These tools are critical in facilitating treatment decisions for each individual patient in an efficient and consistent manner. Due to the often-sparse nature of clinical data, and need to constantly refine treatment decisions, we developed the following decision support workflow for fitting, prediction, and analysis using the ETB framework (**Figure 2A**).

**Figure 2.**
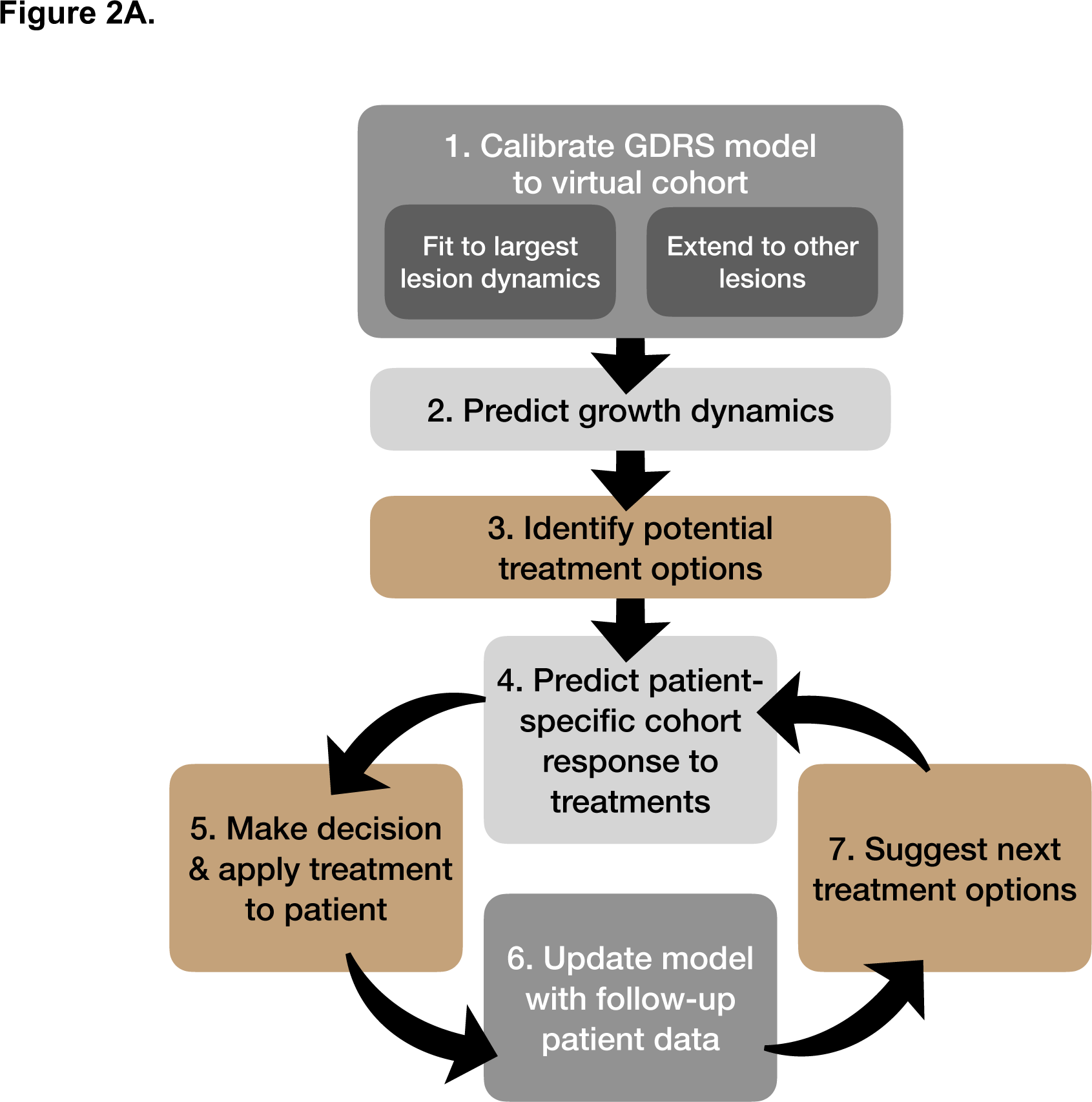
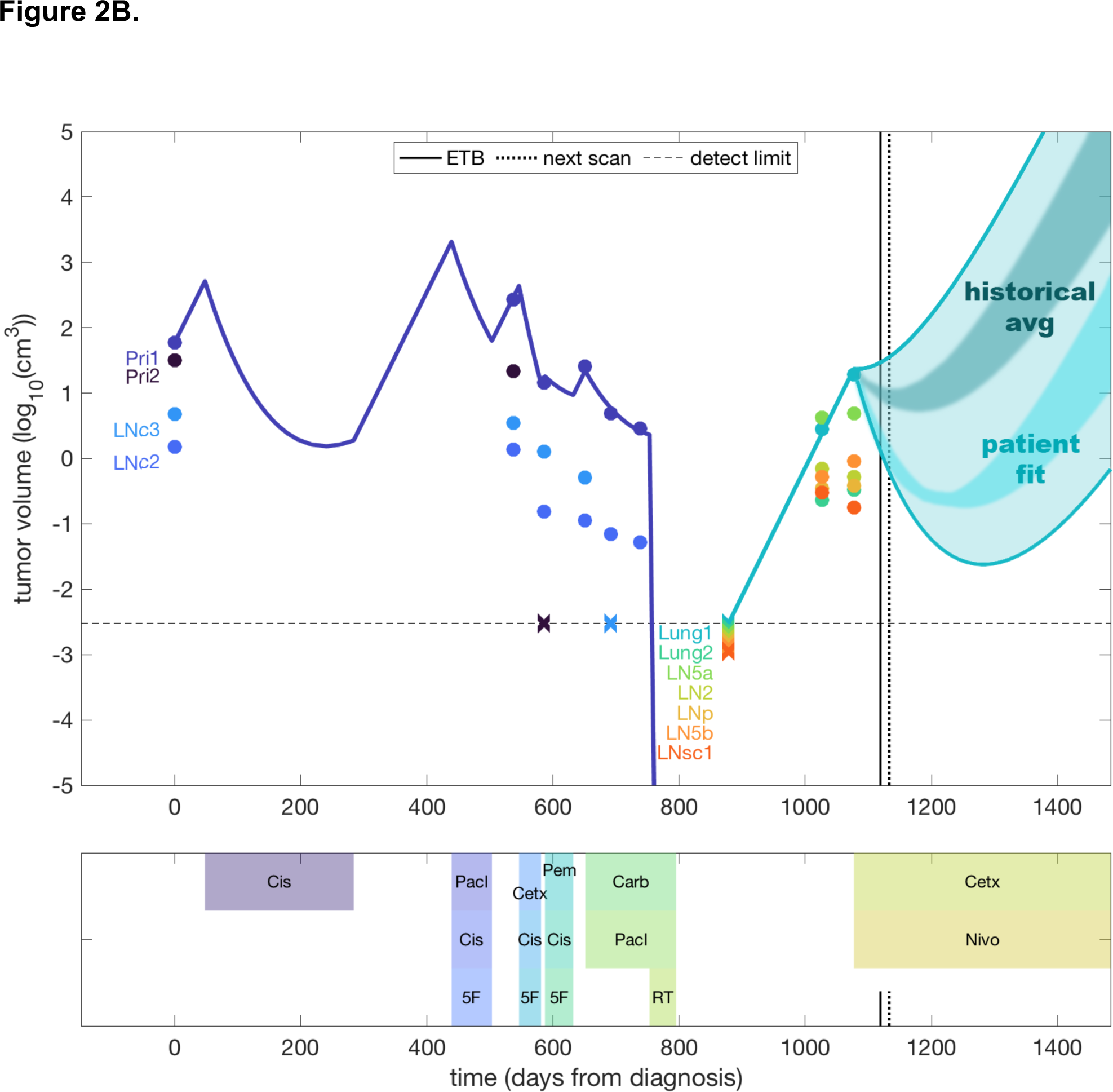
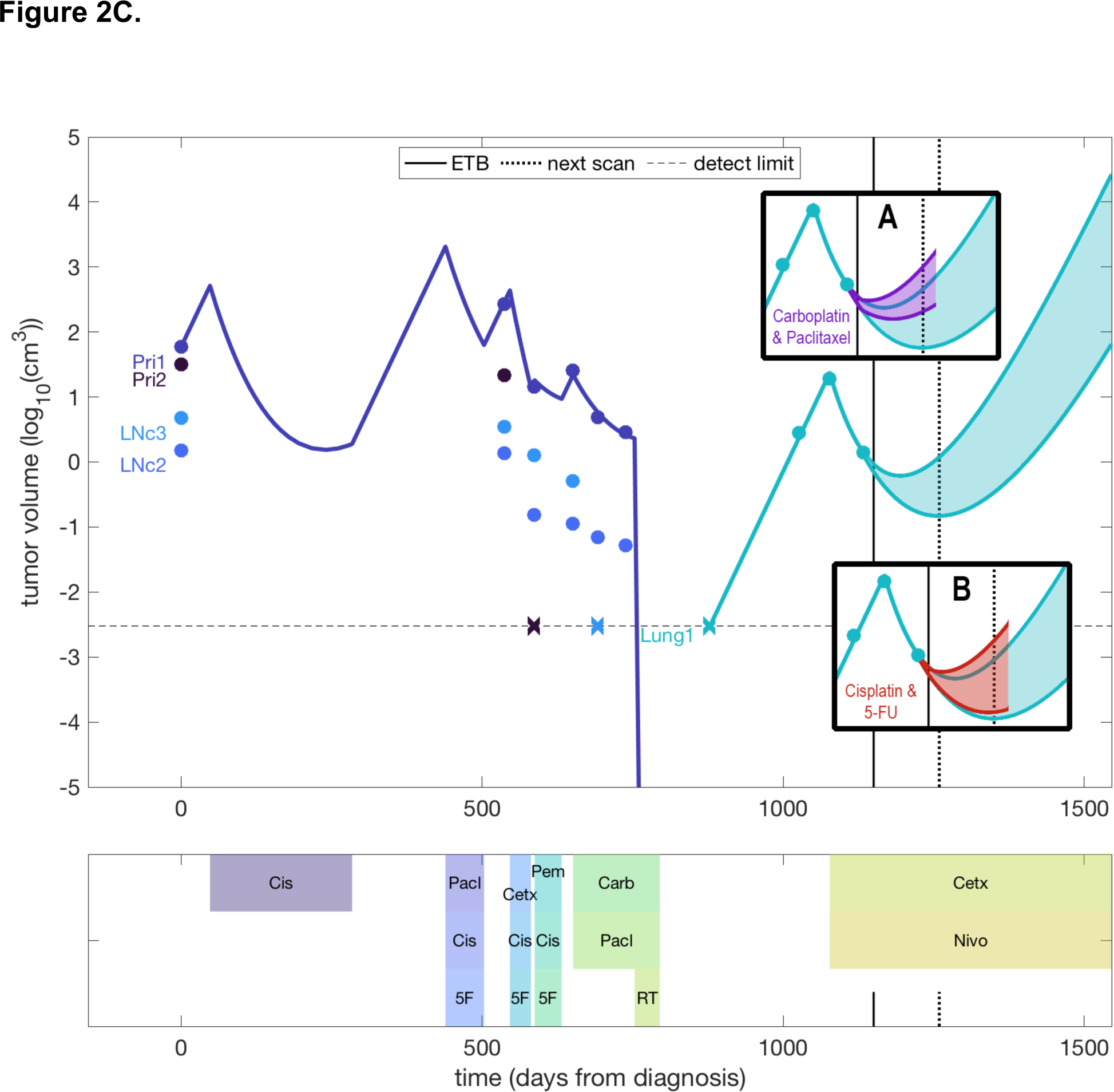
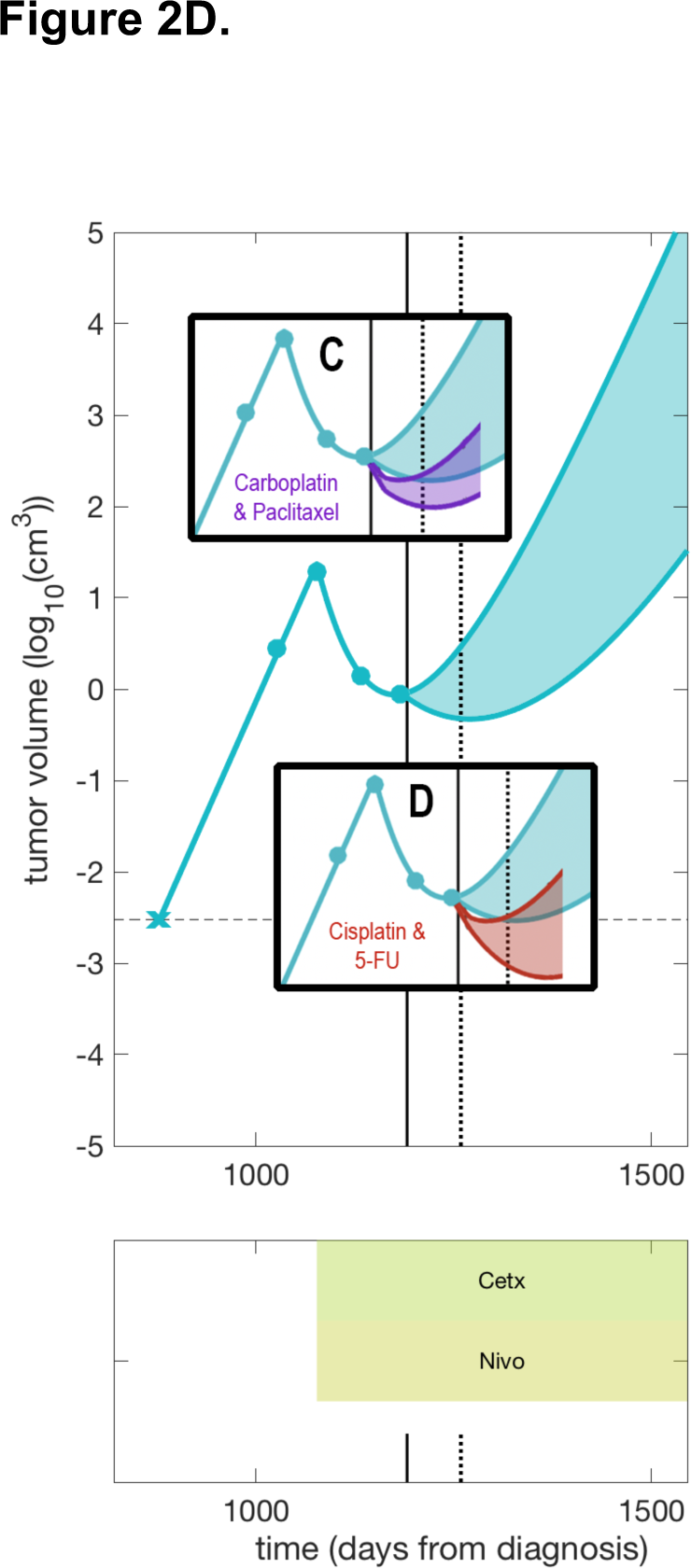
**A:** Overview of the ETB mathematical modeling using the tumor <u>G</u>rowth, tumor <u>D</u>eath, evolution of drug <u>R</u>esistance, and drug re-<u>S</u>ensitization (GDRS) model and decision support workflow. **B:** The initial analysis of the patient. The bottom section of the figure shows the treatments received over time. The top section of plot shows the volumes of each lesion (different colors for each lesion) on a log_10_ scale, over time, with clinical data from imaging represented by dots, and model fits represented by lines. Here, the model fit for the largest primary lesion is plotted (purple), as well as for the largest lung metastasis (teal). The ‘x’ markers represent scans with no detectable volume for that lesion (below the detection threshold for the instrument, represented by the dashed line). The vertical solid line represents the time when the first ETB for the patient occurred, and the vertical dashed line indicated the next expected date of imaging. The model is used to create a cone of outcomes (shaded areas). The wide cone represents the range of outcomes seen in retrospective patients; the inner shaded cones represent the average retrospective patient, and the patient-specific predictions for the current patient. **C:** First follow-up analysis by the ETB (solid vertical line). This occurs shortly after the patient has received their first imaging scan on the current therapy. The new prediction cone is significantly narrowed, given the additional data point. Inset A shows the prediction cone for immediately switching therapy to carboplatin and paclitaxel (purple cone), and inset B shows the predictions if the patient were immediately switched to cisplatin and 5-FU. **D:** Second follow-up analysis by the ETB (solid vertical line). This occurs shortly after the patient’s second scan on current therapy. As in **Figure 2C**, the prediction cones for the two options for second strikes are shown in the insets.

### Evaluation of HNSCC based on the ETB recommendation

To determine the initial feasibility of the ETB based approach, we focused on the evaluation of HNSCC because of the immediate availability of the retrospective cohort through a recently completed clinical trial. Parallel efforts for each enrolled patient in other disease sites to develop a similar model is ongoing and will be reported separately. For HNSCC, we enrolled a man in his 60’s with an initial diagnosis of HNSCC (subject ID: ETB-003) with base of tongue primary site and cervical lymph node metastasis based on imaging studies. The patient pursued non-standard of care alternative therapy and had local disease progression **(Supplemental Table 1)**. The repeat biopsy of cervical lymph node at the time of disease progression was p16-positive squamous cell carcinoma. The patient started palliative chemotherapy with cisplatin, 5-fluorouracil, and cetuximab. Unfortunately, the regimen was discontinued because of toxicity after one cycle. Pembrolizumab was started, and chemotherapy was added due to disease progression on the pembrolizumab monotherapy. Again, the treatment was discontinued due to toxicities. The patient completed the concurrent carboplatin, paclitaxel, and radiation for durable locoregional control. The patient developed disease progression locally and distantly with lung metastasis and was treated with cetuximab and nivolumab.

For this patient with metastatic relapse, we analyzed the potential outcomes that might arise with the application of first-strike second-strike therapy, also known as extinction therapy (**Figure 2B and Figure 2C**). Upon relapse, the patient was put on the combination of cetuximab and nivolumab (the first strike), which we label F1 here. The goal of our analysis was to determine when the patient might fail this combination and therefore intervene at the appropriate time with a second strike. In this case, there were two chemotherapy options available as second strikes: carboplatin plus paclitaxel (S1) and cisplatin plus 5-fluorouracil (S2). Ideally, the first strike will be applied until efficacy wanes and the nadir of tumor burden is near, at which point the switch to the second strike would occur. In our analysis of this patient, we used retrospective cohort data, the patient’s previous imaging data, and the temporal follow-up data to determine 1) when the nadir of F1 may occur, and 2) which of the second strikes to switch to.

### Initial analysis and model fits

**Figure 2B** shows the initial analysis of the patient that was produced after enrollment. The dynamics of their lesions pre-ETB are shown to the left of the solid vertical line, which represents the time at which the patient was first analyzed by the ETB. Some of the early historical data was not available since the patient was treated at another institution prior to being seen at Moffitt Cancer Center. Volumetric measurements for all available scans were performed retrospectively for each detectable lesion and are shown as dots on the plot. The horizontal time axis is scaled relative to the first available scan.

The patient was administered several different therapies to address the primary disease: combination regimens of chemotherapies, targeted therapy, immunotherapy, and radiotherapy, with the latter causing regression of the primary disease around day 760. A follow-up scan on day 878 showed no evidence of disease; however, on day 1026 there was evidence of lung and lymph node metastases. These were measured volumetrically. Knowing their positions in the lungs, the previous scan with no evidence of disease (NED) at day 878 was reexamined to see if very small lesions were indeed detectable, but they remained NED. Therefore, we consider that these lesions are smaller than the detection threshold of the instrument, and they are marked with ‘x’ markers on **Figure 2B**. Shortly before enrolling on the ETB, the patient began their first-strike therapy (F1) for the metastatic disease.

In accordance with the methods, we fit the model to the available data. The growth rate ranges were primarily fit from the metastatic disease dynamics, since there were two measurements prior to starting therapy F1, and an additional upper limit from the NED scan on day 878. Using these ranges of growth rate, we fit efficacy and resistance parameters for the dynamics of the primary disease (of which a representative fit for the largest primary lesion is shown in **Figure 2B** in dark blue). A confounding factor is that the drugs were primarily given in combination, and furthermore the imaging data is sparse compared to the multiple changes of agents. However, some constraints on the drug behaviors can be gained from these fits.

We also leveraged our retrospective data for the first-strike therapy from patients having received the same F1 combination of cetuximab and nivolumab, from the clinical trial described above. By fitting to the dynamics of the patients in that cohort, we determined ranges of efficacy for F1 (**Table 2**). Application of this range to the current patient (using the intrinsic range of growth rates and resistance rates found from their own lesion dynamics) produced the predictive cone shown in light blue (the widest cone). Naturally, since some retrospective patients progressed rapidly and others had significant responses, the cone encompasses a wide range of possible responses for the current patient. Taking the average retrospective behavior and applying it to the current patient produces the darker shaded region.

At this stage of the analysis and patient decision-making process, we are primarily interested in knowing when the efficacy of F1 will be significantly diminished, and therefore the nadir of tumor volume will be approached. This will be the time to switch to a second strike. The time-to-nadir (TTN) for the retrospective cohort parameter ranges applied broadly to this patient ranges from 0 months (i.e., the efficacy of F1 is low enough that the lesions are already growing through it) to 7.3 months. To refine the patient-specific prediction of the nadir, we leverage model fits derived from the earlier lesion dynamics, and restrict the fits generated by the retrospective cohort parameter ranges to those that match the current patient’s fits. After this constraint, the model predicts that the current patient is likely to do significantly better than the average response of the retrospective cohort. The likely TTN range is now between 4.2 months and 6.9 months. Since the next scan is anticipated to be within two months of starting the therapy, the model strongly suggests that switching therapies should wait until follow-up imaging is obtained.

### First follow-up analysis

Upon imaging and performing volumetric measurements of the lesions, we reanalyze the patient dynamics. **Figure 2C** shows the results after the first follow-up scan for the patient at day 1119. The largest lesion has declined significantly under the first strike, F1. This decline was in line with the “Patient fit” prediction cone of **Figure 2B**, suggesting that the growth rates and treatment dynamics determined from earlier timepoints remained consistent for this lesion over time. The updated prediction cone for F1 is narrower after follow-up analysis using the additional data point. The TTN now ranges from 1.5 to 3.5 months (from the time of follow-up, not the start of F1). The model again suggests that the first strike is most likely to remain efficacious until the next imaging cycle, with only a small fraction of simulations suggesting that the nadir will be reached prior to that time.

At the same time, we examine what the effect of switching to a second strike would be at this time, since we do not want to wait until the nadir is reached to switch. In the insets of **Figure 2C**, we show the predicted range of effect for switching from the F1 to either second-strike S1 (inset A) or S2 (inset B). In both cases, the range of efficacy for the strike is determined by both retrospective cohort responses and the current patient’s response, since they were previously administered these agents during primary tumor treatment. S1 was administered at the end of the primary disease treatment, and therefore the estimates are better than for S2, which was only administered in combination with other agents, and therefore has confounding factors in the primary fits. However, in both cases, the model suggests that compared to staying with the first strike, the second strikes appear to bring no advantage at this time. Therefore, since the model predicts that there is likely some efficacy remaining in F1 and that both S1 and S2 currently provide little comparative advantage, the decision is to continue the therapy until the next imaging time point.

### Second follow-up analysis

For any patient in follow-up, the ETB process repeats with each new scan. Volumetric measures for the next follow-up scan were attained and the updated analysis is shown in **Figure 2D**. The tumor has continued to shrink, albeit at a slower rate than during the initial phase. Reanalyzing the data with the model leads to updated predictions for the TTN, which now ranges from 0 to 1.7 months (from the reanalysis time point). This suggests that the efficacy of F1 is approaching its end. The insets in the figure show the predicted efficacy of the two second-strike options, and as opposed to **Figure 2C**, both now are likely to have a better effect on tumor burden than continuing with F1.

### Decision support in the ETB

The ETB is a non-interventional trial, and therefore the decisions of when to switch therapies and what to switch to remain in the hands of the oncologist and the patient. Here, the model analysis and predictions used in the ETB workflow aim to give insight into the temporal dynamics of the patient’s specific disease, which can aid in making the above decisions. In the exemplar case above, the model initially suggested continuing the first strike, and similarly continuing after the first follow-up imaging point, after which the model began to suggest that a second-strike option should be applied soon. For this patient, an additional treatment option, radiotherapy, arose at the time, and was chosen as a second strike. The insight gained regarding the efficacy and expected nadir of the first strike was valuable in making the subsequent treatment decisions for the patient.

## DISCUSSION

Here we report the first results of the ETB feasibility study, having built a system to capture available clinical tumor dynamics over time and in response to available therapies, to incorporate the remaining conventional therapeutic options, and created a forum and methodology for disciplines with an evolutionary understanding of cancer to discuss ideas and implement them for patients. We also highlight a specific patient to provide more detail of the process of building a model and using it to guide timing and selection of available therapies in a novel and dynamic manner. We leveraged the existing framework of clinical tumor boards, the well-established best practice care delivery model in oncology, in combination with quantitative analysis^7,15^. ETB uniquely includes non-clinical disciplines and integrates eco-evolutionary concepts to generate novel treatment strategies and assesses the results of this platform over time.

To date we have primarily analyzed patients with HNSCC while additional investigations into adolescent and young adult sarcomas, prostate, breast, and lung cancers are ongoing. Successfully expanding to additional disease sites depends on many factors. A key aspect is having the option to change therapies in real time during follow-up. In addition, availability of retrospective cohorts to inform model development is critical to their utility. **Figure 3** shows how we envision the current incarnation of the ETB in facilitating the integration of evolutionary therapies into standard practice. Early on, the ETB serves as a test bed for developing models, understanding the evolutionary dynamics of the disease, and creating potential clinical trials. In later stages, pilot trials will be served by a robust framework using highly calibrated mathematical model(s) developed for that trial. If successful, we envision developing disease-specific software modules that will assist the oncologist in decision support during standard of care, particularly in practice environments without access to multidisciplinary tumor board. This long-term vision delivers the insights gained from mathematical modeling of evolutionary dynamics into the hands of any interested oncologist for personalized decision support.

**Figure 3.**
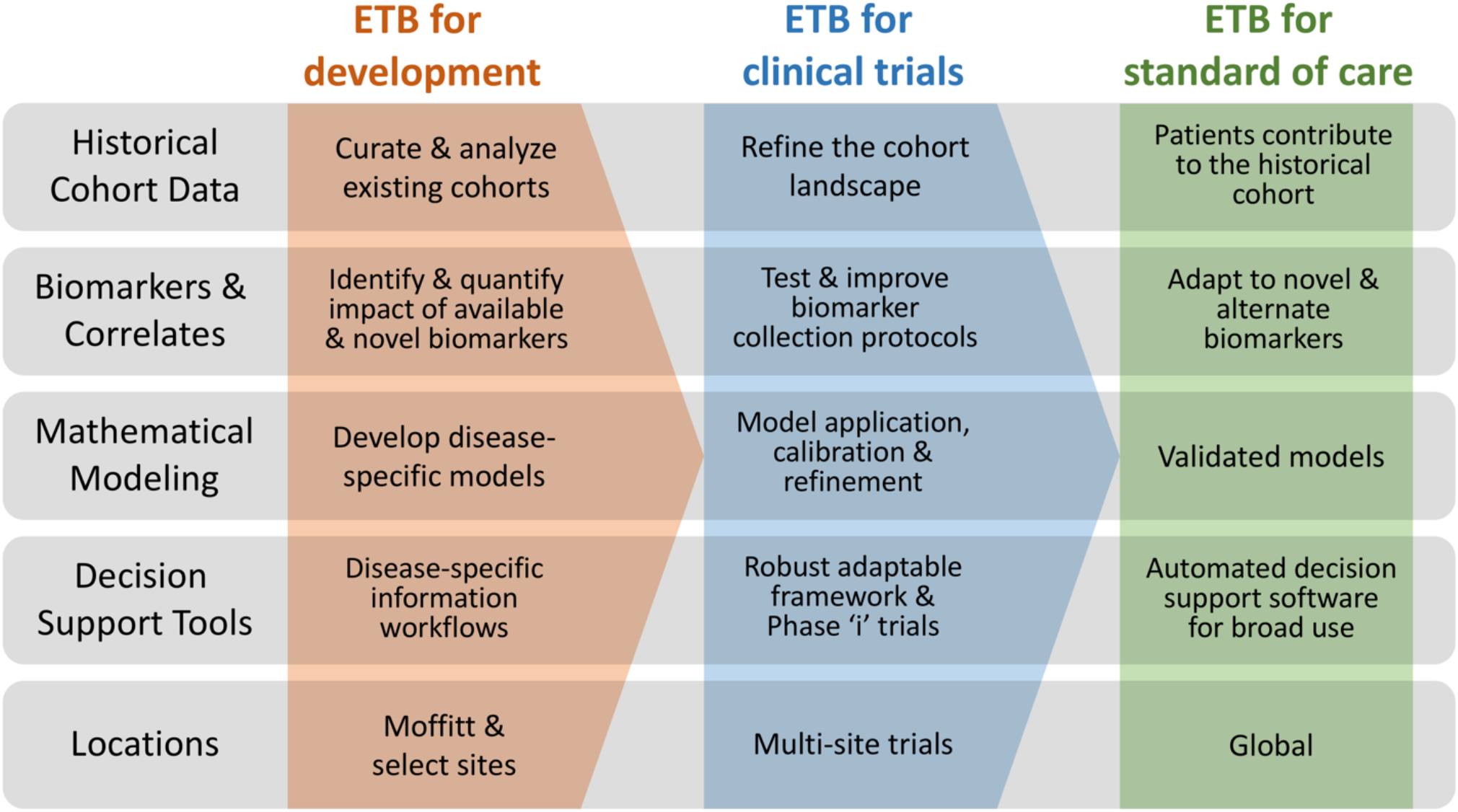
Three phases of the ETB. The development phase (red) is where models and evolutionary therapy approaches are developed for each specific disease setting based on available biomarkers. Retrospective cohorts are curated and analyzed. The second phase (blue) brings the ETB approach to disease-specific clinical trials, using a Phase ‘i’ virtual trial approach for patient decision support. Models and biomarker collection are refined, and patient cohorts are expanded. In phase three (green), validated approaches are deployed for broad clinical use, via custom software developed for the specific disease. This system can be self-improving with each patient that is seen via the approach, contributing valuable data and outcomes to the retrospective cohort for a given disease.

The ETB invites consideration of evolutionarily enlightened therapy approaches. Our primary focus here has been using multi-strike therapy to effect maximal tumor reduction, ideally extinction, while minimizing the effect of tumor resistance to any given agent. Evolutionary theory suggests that it is best to switch therapies either just before or at the nadir of the tumor volume under the current therapy (as opposed to waiting for progression to occur and be observed on imaging) because the nadir is believed to represent the point that the tumor population has effectively evolved resistance to the first-strike therapy. A key tool in this approach is predictive modeling, which can estimate time-to-progression before it occurs. In the best case, multi-strike therapy can effect a cure, but even in the absence of such, the approach has the potential to deliver deeper remission, both in terms of tumor volume and PFS.

There are several other eco-evolutionary concepts that are already in application or in development. One of the more developed is adaptive therapy, which typically exploits the evolutionary costs of resistance such as synthesis, maintenance, and operation of the molecular machinery needed to survive treatment. The benefits of resistance exceed costs during therapy. This has best been demonstrated thus far clinically in prostate cancer^16^ but also in preclinical models of breast cancer^17^. Adaptive therapy is predicted to be most effective when there is a competitively dominant sensitive population of cancer cells, typically present in clinical situations that have high response rate but a low cure rate. It also requires a treatment strategy that reliably induces a response. With continuous therapy or metronomic therapies that do not consider the patient’s tumor dynamics, competitively subordinate resistant cancer cells come to predominate as they experience competitive release from the absence of the drug-sensitive cancer cells^18-20^. By removing therapy while there is still a high frequency and density of sensitive cells, these sensitive cells effectively compete with and therefore suppress the resistant cancer cells. Adaptive therapy is an important and active area of evolutionary therapy research^16,17,19,21-32^ that can encompass multiple drugs^31,32^ and has already been considered for some ETB patients.

We plan to initiate several clinical trials to better examine the ETB approach with respect to accuracy, predictability, and outcomes. However, much work remains to be done. First, the models will be improved with more real-world data, and thus increased analysis of additional retrospective cohorts in each disease will serve to better constrain the operating parameter ranges we apply in our models. Second, compared to imaging, collecting additional biomarkers such as cfDNA has the potential to provide faster turnaround, more frequent and even potentially more accurate information on tumor dynamics, particularly in minimal residual disease states. Third, streamlining the process of clinical annotation, literature searches, biomarker collection, analysis, and prediction will be necessary to apply this labor-intensive workflow on a wider scale, though having performed the process for a given diagnosis provides efficiencies for future ETB patients. To this end, we are developing a framework that automates much of the process from start to finish, allowing the ETB team to focus on key discussion points regarding the model predictions and patient assumptions that drive the next decision point. Ultimately, we hope to continue to improve the therapeutic paradigm of clinical oncology application based on the insight gained from ETB. We anticipate the tools generated will inform both sequences and strategies of multi-strike therapy across a number of cancers and directly facilitate the likelihood of cancer extinction.

## Supporting information

Supplemental Documentation

## Data Availability

Most data produced in the present work are contained in the manuscript. All data produced in the present study are available upon reasonable request to the authors.

## Notes

**Potential Conflict of Interest:** HE holds U.S. Patent 63/279,994: Bayesian Framework to Augment Tumor Board Decision Making (provisional), U.S. Patent 62/040,579: Predicting glioma treatment response (provisional), and U.S. Patent 62/944,804: Methods for prostate cancer intermittent adaptive therapy (provisional). TB received Research funding support from Bristol Myers Squibb and has stock ownership in Ionis Pharmaceuticals. HS received honoraria from AstraZeneca, Seattle Genetics, Merck, and Novartis for serving in *ad hoc* scientific advisory boards. DRR received honoraria from Springworks and Eisai for serving in Data Safety Monitoring Committee. CHC received honoraria from Sanofi, Merck, Brooklyn ImmunoTherapeutic, Fulgent, and Exelixis for serving in *ad hoc* scientific advisory boards. All other authors declared no conflict of interest.

### Competing Interest Statement

HE holds U.S. Patent 63/279,994: Bayesian Framework to Augment Tumor Board Decision Making (provisional), U.S. Patent 62/040,579: Predicting glioma treatment response (provisional), and U.S. Patent 62/944,804: Methods for prostate cancer intermittent adaptive therapy (provisional). TB received Research funding support from Bristol Myers Squibb and has stock ownership in Ionis Pharmaceuticals. HS received honoraria from AstraZeneca, Seattle Genetics, Merck, and Novartis for serving in ad hoc scientific advisory boards. DRR received honoraria from Springworks and Eisai for serving in Data Safety Monitoring Committee. CHC received honoraria from Sanofi, Merck, Brooklyn ImmunoTherapeutic, Fulgent, and Exelixis for serving in ad hoc scientific advisory boards. All other authors declared no conflict of interest.

### Clinical Protocols

https://clinicaltrials.gov/ct2/show/NCT04343365

### Funding Statement

This study was funded by Moffitt Cancer Center and the Center of Excellence for Evolutionary Therapy at Moffitt Cancer Center

### Author Declarations

Protocol No.: 20417 Title: Feasibility of Generating Novel Therapeutic Strategies Based on Evolutionary Tumor Board in Cancer Patients Protocol Status.: IRB INITIAL APPROVAL PI: Chung, Christine Institution: Moffitt Cancer Center IRB No.: IRB Committee: Advarra Meeting Date: 04/07/2020 Review Reason: Initial Review Review Type: Expedited Action: Approved Action Effective Date: 04/07/2020 Action Expiry Date: 04/07/2021

## REFERENCES

1 Siegel, R. L., Miller, K. D., Fuchs, H. E. & Jemal, A. Cancer statistics, 2022. CA Cancer J Clin 72, 7–33 (2022). https://doi.org:10.3322/caac.21708

2 Stankova, K., Brown, J. S., Dalton, W. S. & Gatenby, R. A. Optimizing Cancer Treatment Using Game Theory: A Review. JAMA Oncol 5, 96–103 (2019). https://doi.org:10.1001/jamaoncol.2018.3395

3 Gatenby, R. A. & Brown, J. Mutations, evolution and the central role of a self-defined fitness function in the initiation and progression of cancer. Biochim Biophys Acta 1867, 162–166 (2017). https://doi.org:10.1016/j.bbcan.2017.03.005

4 Gravenmier, C. A., Siddique, M. & Gatenby, R. A. Adaptation to Stochastic Temporal Variations in Intratumoral Blood Flow: The Warburg Effect as a Bet Hedging Strategy. Bull Math Biol 80, 954–970 (2018). https://doi.org:10.1007/s11538-017-0261-x

5 Gatenby, R. A., Zhang, J. & Brown, J. S. First Strike-Second Strike Strategies in Metastatic Cancer: Lessons from the Evolutionary Dynamics of Extinction. Cancer Res (2019). https://doi.org:10.1158/0008-5472.CAN-19-0807

6 Gatenby, R. A., Artzy-Randrup, Y., Epstein, T., Reed, D. R. & Brown, J. S. Eradicating metastatic cancer and the eco-evolutionary dynamics of Anthropocene extinctions. Cancer Res (2019). https://doi.org:10.1158/0008-5472.CAN-19-1941

7 Kim, E., Rebecca, V. W., Smalley, K. S. & Anderson, A. R. Phase i trials in melanoma: A framework to translate preclinical findings to the clinic. Eur J Cancer 67, 213–222 (2016). https://doi.org:10.1016/j.ejca.2016.07.024

8 Chung, C. H. et al. Concurrent Cetuximab and Nivolumab as a Second-Line or beyond Treatment of Patients with Recurrent and/or Metastatic Head and Neck Squamous Cell Carcinoma: Results of Phase I/II Study. Cancers 13 (2021). https://doi.org:10.3390/cancers13051180

9 Chung, C. H. et al. Phase II multi-institutional clinical trial result of concurrent cetuximab and nivolumab in recurrent and/or metastatic head and neck squamous cell carcinoma. Clin Cancer Res (2022). https://doi.org:10.1158/1078-0432.CCR-21-3849

10 Brady, R. & Enderling, H. Mathematical Models of Cancer: When to Predict Novel Therapies, and When Not to. Bull Math Biol 81, 3722–3731 (2019). https://doi.org:10.1007/s11538-019-00640-x

11 Claret, L. et al. Model-based prediction of phase III overall survival in colorectal cancer on the basis of phase II tumor dynamics. J Clin Oncol 27, 4103–4108 (2009). https://doi.org:10.1200/JCO.2008.21.0807

12 Claret, L. et al. Evaluation of tumor-size response metrics to predict overall survival in Western and Chinese patients with first-line metastatic colorectal cancer. J Clin Oncol 31, 2110–2114 (2013). https://doi.org:10.1200/JCO.2012.45.0973

13 Glazar, D. J. et al. Tumor Volume Dynamics as an Early Biomarker for Patient-Specific Evolution of Resistance and Progression in Recurrent High-Grade Glioma. J Clin Med 9 (2020). https://doi.org:10.3390/jcm9072019

14 Glazar, D. J. et al. Early response dynamics predict treatment failure in patients with recurrent and/or metastatic head and neck squamous cell carcinoma treated with cetuximab and nivolumab. Oral Oncol 127, 105787 (2022). https://doi.org:10.1016/j.oraloncology.2022.105787

15 Pasetto, S., Gatenby, R. A. & Enderling, H. Bayesian Framework to Augment Tumor Board Decision Making. JCO Clin Cancer Inform 5, 508–517 (2021). https://doi.org:10.1200/CCI.20.00085

16 Zhang, J., Cunningham, J. J., Brown, J. S. & Gatenby, R. A. Integrating evolutionary dynamics into treatment of metastatic castrate-resistant prostate cancer. Nat Commun 8, 1816 (2017). https://doi.org:10.1038/s41467-017-01968-5

17 Enriquez-Navas, P. M. et al. Exploiting evolutionary principles to prolong tumor control in preclinical models of breast cancer. Sci Transl Med 8, 327ra324 (2016). https://doi.org:10.1126/scitranslmed.aad7842

18 Gillies, R. J., Flowers, C. I., Drukteinis, J. S. & Gatenby, R. A. A unifying theory of carcinogenesis, and why targeted therapy doesn’t work. Eur J Radiol 81 Suppl 1, S48–50 (2012). https://doi.org:10.1016/S0720-048X(12)70018-9

19 Gatenby, R. A., Silva, A. S., Gillies, R. J. & Frieden, B. R. Adaptive therapy. Cancer Res 69, 4894–4903 (2009). https://doi.org:10.1158/0008-5472.CAN-08-3658

20 Gatenby, R. A., Brown, J. & Vincent, T. Lessons from applied ecology: cancer control using an evolutionary double bind. Cancer Res 69, 7499–7502 (2009). https://doi.org:10.1158/0008-5472.CAN-09-1354

21 Brady-Nicholls, R. et al. Prostate-specific antigen dynamics predict individual responses to intermittent androgen deprivation. Nat Commun 11, 1750 (2020). https://doi.org:10.1038/s41467-020-15424-4

22 Brady-Nicholls, R. et al. Predicting patient-specific response to adaptive therapy in metastatic castration-resistant prostate cancer using prostate-specific antigen dynamics. Neoplasia 23, 851–858 (2021). https://doi.org:10.1016/j.neo.2021.06.013

23 Cunningham, J. et al. Optimal control to reach eco-evolutionary stability in metastatic castrate-resistant prostate cancer. PLoS One 15, e0243386 (2020). https://doi.org:10.1371/journal.pone.0243386

24 Gallaher, J. A., Enriquez-Navas, P. M., Luddy, K. A., Gatenby, R. A. & Anderson, A. R. A. Spatial Heterogeneity and Evolutionary Dynamics Modulate Time to Recurrence in Continuous and Adaptive Cancer Therapies. Cancer Res 78, 2127–2139 (2018). https://doi.org:10.1158/0008-5472.CAN-17-2649

25 Kim, E., Brown, J. S., Eroglu, Z. & Anderson, A. R. A. Understanding the potential benefits of adaptive therapy for metastatic melanoma. bioRxiv, 2020.2010.2016.343269 (2020). https://doi.org:10.1101/2020.10.16.343269

26 Kim, E., Brown, J. S., Eroglu, Z. & Anderson, A. R. A. Adaptive Therapy for Metastatic Melanoma: Predictions from Patient Calibrated Mathematical Models. Cancers (Basel) 13 (2021). https://doi.org:10.3390/cancers13040823

27 Mason, N. T. et al. Budget Impact of Adaptive Abiraterone Therapy for Castration-Resistant Prostate Cancer. Am Health Drug Benefits 14, 15–20 (2021).

28 Strobl, M. A. R. et al. Spatial structure impacts adaptive therapy by shaping intra-tumoral competition. bioRxiv, 2020.2011.2003.365163 (2021). https://doi.org:10.1101/2020.11.03.365163

29 Strobl, M. A. R. et al. Turnover Modulates the Need for a Cost of Resistance in Adaptive Therapy. Cancer Res 81, 1135–1147 (2021). https://doi.org:10.1158/0008-5472.CAN-20-0806

30 West, J., Ma, Y. & Newton, P. K. Capitalizing on competition: An evolutionary model of competitive release in metastatic castration resistant prostate cancer treatment. J Theor Biol 455, 249–260 (2018). https://doi.org:10.1016/j.jtbi.2018.07.028

31 West, J. et al. Towards Multidrug Adaptive Therapy. Cancer Res 80, 1578–1589 (2020). https://doi.org:10.1158/0008-5472.CAN-19-2669

32 West, J. B. et al. Multidrug Cancer Therapy in Metastatic Castrate-Resistant Prostate Cancer: An Evolution-Based Strategy. Clin Cancer Res (2019). https://doi.org:10.1158/1078-0432.CCR-19-0006

