## Supplemental Documentation for "Feasibility of an Evolutionary Tumor Board for Generating Novel Personalized Therapeutic Strategies"

**Supplemental Table 1. Example of prior therapy data collection form from subject ETB-003.**

| Regimen | Reference | Cycle Length (days) | # Cycles | Best Response | Known Resistance Mechanism | ETB Comment |
|---|---|---|---|---|---|---|
| Insulin potentiation therapy (IPT) - Cisplatin | PMID: 30712796 | N/A | 2 | N/A | N/A | IPT is not a proven anticancer treatment |
| Insulin potentiation therapy (IPT) – paclitaxel + cisplatin + 5-fluorouracil | PMID: 30712796 | N/A | N/A | N/A | N/A | IPT is not a proven anticancer treatment |
| cisplatin + 5-fluorouracil + cetuximab | PMID: 18784101 | 21 | 2 | PR | Tumor phenotype change to mesenchymal phenotype | Initiated with cisplatin, switched to carboplatin due to elevated creatinine. Discontinued regimen due to toxicity. |
| cisplatin + 5-fluorouracil + pembrolizumab | PMID: 31679945 | 21 | 3 | PR | Downregulation of antigen presenting machinery | Initiated as pembrolizumab alone, added chemo due to visible growth. Discontinued pembrolizumab after 2 doses due to possible immune-mediated toxicity. Switched to carboplatin again due to elevated creatinine |
| carboplatin + paclitaxel | PMID: 29774120 | 21 | 4 | CR | N/A | |
| carboplatin + paclitaxel + radiation therapy | PMID: 14645636 | 30 | 1 | CR | TP53 mutation, low tumor infiltrating lymphocyte, tobacco use history | 6,600 cGy IMRT/IGRT over 30 fractions, given concurrent with carboplatin + paclitaxel regimen for locoregional control |

N/A: Not Available, PR: Partial Response, CR: Complete Response

**Supplemental Table 2. Example of available therapies and anticipated outcomes**

| Regimen | Published Complete Response Rate | Published Event-Free Survival | Published Median Overall Survival | Reference (e.g. PMID) |
|---|---|---|---|---|
| Docetaxel | 5.4% | 6.5 months median duration of response | Not reported | PMID: 7918125 |
| Methotrexate | 0.7% | Not reported | 6.7 months | PMID: 19289630 |
| Nivolumab/Cetuximab | Not available | Not available | Not available | NCT03370276 |
| Carboplatin/Paclitaxel | 4% | 4.7 months median progression-free survival | 9.1 months | PMID: 29774120 |

**Supplemental File 1**

**Development of Therapeutic Strategies based on Mathematical Models**

The growth of each lesion (each individual metastasis) is modeled using exponential growth with parameter $\gamma_i$. Tumor growth rates are not always perfectly exponential, particularly at larger sizes; we favored this growth model because using a more complex growth law would increase the number of parameters with minimal benefit to fitting the patient data. To date, we have found that the exponential growth model provides an excellent fit across lesions of different sizes both within and across patients, for several different cancers. This superior fit relative to other growth models is likely due to the paucity of data points.

The reduction in lesion volumes caused by each drug is also modeled as an exponential term. In effect, each drug causes a reduction of the growth rate, and in most cases, for highly cytotoxic drugs, the death term will be larger than the growth term, causing a decline in the size of that lesion. For a single drug *j*, the product of the maximal drug death rate ($\delta_j$), the current efficacy ($E_j \in [0,1]$), and the current dose ($D_j \in [0,1]$) determines the ecological dynamics of the tumor when the drug is on ($D_j > 0$). If that product is greater than $\gamma_i$, then the lesion will decrease in size. If the product is less than $\gamma_i$, then the lesion will grow, but at a slower rate than the untreated tumor.

The evolution of cancer cells within each tumor is modelled by the time-dependent drug efficacy term $E_j$. We assume that the efficacy of a drug decays exponentially while the drug is on (representing the progressive selection of drug resistance in the tumor), and that the efficacy increases logistically toward 1 when the drug is not being administered (representing the re-sensitization to a previous therapy). Note that the resensitization dynamic is only relevant to the model for cases where the tumor(s) are rechallenged with a previously used therapy.

The GDRS model was developed from serial observations of volumetric measurements of individual lesions. Time series data came from retrospective cohorts as well as the patients enrolled on the ETB protocol. Namely, we often found the pattern of an initial decline in tumor size when a drug was commenced, followed by decreasing efficacy and eventual regrowth. The GDRS model generally provided excellent fits to these common ‘U-shaped’ dynamics. This fit is accomplished with a minimum number of parameters (1 growth parameter, and 2 parameters per drug). Our overall approach to the ETB is fully flexible and can use alternative models of tumor growth and treatment, but here we focus on the results produced with the GDRS model.

**Parameter space mapping from patient data**

In the exemplar patient of Figures 2B-D, the model plot prior to the ETB line is one “fit” out of many possible trajectories. For simplicity, we show one trajectory but a range of model fits match this data. To find exactly where these fits are in parameter space, our approach was as follows. First, we fit a growth rate from the data. In the exemplar patient, we used a growth rate corresponding to the largest metastatic lesion at the time of the ETB. This lesion had two pre-treatment imaging points, which gave a growth rate of 0.038. The scan before detection showed no apparent lesion, and applying our assumptions about detection size, we find that this growth rate is also consistent with the lesion being present but below the detection level of the instrument.

Once the growth rate was selected, we defined the parameter space of death and resistance for each segment of therapy (segment *j*). Note that resensitization only comes into play upon repeat application of therapies, and therefore is not part of the initial fitting for death and resistance. In all cases, there is an imaging point prior to the start of therapy *j*. Furthermore, there may be one or more imaging points following therapy before the next therapy begins. In general, the final imaging point after starting therapy *j* becomes the pre-treatment value for the next segment. For the case where there is one pre-treatment point and more than one on-treatment point, we can generally find a set of parameters $\{\delta_j, r_j\}$ that fits the three points. The exception being if the points are concave down, in which case the model is poorly defined. For the case where there is only one imaging point on treatment, then there is a space of $\{\delta_j, r_j\}$ that would go through pre- and on-treatment points. One endpoint of that space is where $r_j = 0$ and $\delta_j$ fits the constant exponential change from pre- to on- treatment points. With $r_j > 0$, solving the GDRS model equations leads to the corresponding fitting value for $\delta_j$, given by:

$$\delta_j = \frac{r_j\left(\gamma_j \Delta t - \ln(T_1/T_0)\right)}{1 - e^{-r_j \Delta t}}$$

where $T_1$ is the on-treatment volume, $T_0$ is the volume at start of therapy, and $\Delta t$ is the time between the two scans. In the limit as $r_j \rightarrow 0$, the death parameter is a straight forward solution of exponential decline, as long as $\delta_j > \gamma_j$ :

$\delta_j = \gamma_j - \frac{\ln(T_1/T_0)}{\Delta t}$.

The upper endpoint of the space of fitting {$\delta_j$, $r_j$} is essentially unlimited since any value of $r_j$ can be matched by a corresponding value of $\delta_j$. In practice, there are limits to this fit. First, real tumors that decline below one cell would be removed and would not regrow, so this puts a limit on the depth of the "u-shape" and therefore an upper limit on the space of fitting {$\delta_j$, $r_j$}. In practice, we also limit the range of $\delta_j$ based on values observed in the historical cohort, as described in Table 2. This procedure leads to bounds on the space of {$\delta_j$, $r_j$} for a given growth rate, and the formation of prediction cones and the average trajectory applies the extremes of this space to the patient in a predictive fashion. In Figure 2B, the patient-specific trajectory part of the cone arises due to the fact that previous administration of these agents in that patient led to patient-specific estimates for $\delta$ and $r$ for the cetuximab and nivolumab.

Combinations of agents can be treated in two ways. If none of the agents are used again, then the combination can be considered as a single therapy, because the individual contributions of each agent to the dynamics do not have importance. However, when there are multiple agents used and one or more are later applied separately (as in the case of cetuximab for the exemplar patient), we also sweep the contributions of each agent in that initial combination. In this case, resensitization also becomes a factor. In this study, we have set the resensitization to be equal to the rate of resistance. In this case, the change of contribution to one agent can affect subsequent fits of later strikes. However, these efficacies contribute to overall values of {$\delta$, $r$} for the combination as a whole; the individual efficacies therefore can be adjusted algebraically across the fitting parameters. For example, if drugs A and B are applied early and then drugs B and C are applied later, changes to the efficacy parameter of drug B in the first combination are also propagated to the second combination, and then the efficacy of drug C is adjusted to compensate for the change in B.

In this example, we have used a basic approach to fitting the data to find average and extreme behaviors of a historical cohort and applied that to patient specific measures to produce predictive cones. More sophisticated methods might include forming distributions of parameters from historical data, and weighting fits by error measures such as least squares or similar. However, given the sparsity of the patient data and the range of dynamics possible, for our purposes of constraining and comparing the dynamics of various treatments we chose an approach that was parsimonious. Current and future work will include larger historical datasets that are analyzed, propagated, and illustrated with more statistical-based methods.
